# SCOPING REVIEW PROTOCOL OF E-HEALTH INTERVENTIONS TARGETING INDIGENOUS POPULATIONS

**DOI:** 10.1101/2025.10.16.25338190

**Authors:** Shikha Shah, Ginny Monteiro, Ronak Borana, Tyra Yarran, Stephanie Topp, Maria Eugenia Castellanos

**Affiliations:** Rollins School of Public Health, Emory University, Atlanta, USA; Public Health and Tropical Medicine, College of Medicine and Dentistry, James Cook University, Townsville, Queensland, Australia; Cape York Institute for Policy & Leadership, Australia; XLRI Jamshedpur, India; North Metropolitan Health Service - Aboriginal Health, Health Department Western Australia, Australia

**Keywords:** Indigenous Peoples, Health Services, Indigenous, Telemedicine, Mobile Applications

## Abstract

**Introduction:** Electronic Health (E-Health) technologies present a valuable opportunity to enhance healthcare access and outcomes for Indigenous populations by addressing persistent, often structurally determined, health disparities. Despite increasing recognition of the potential of E-Health interventions, there remains a lack of comprehensive synthesis of their modalities, cultural appropriateness, and implementation within Indigenous communities globally. This scoping review aims to systematically map the landscape of E-Health interventions for Indigenous populations, identifying effective strategies and gaps in current research and practice. By documenting global evidence, this review contributes to broader efforts to integrate Indigenous health considerations into policy and practice, supporting health equity and realising the United Nations Declaration on the Rights of Indigenous Peoples.

**Methods and Analysis:** This scoping review will follow the Preferred Reporting Items for Systematic Reviews and Meta-Analysis Protocols (PRISMA-P) Guidelines. A comprehensive search of multiple databases will be conducted to identify studies on E-Health interventions in Indigenous communities between 2013 and 2024. The review will focus on interventions targeting Indigenous populations published in English. A two-stage screening process, comprising abstract/title screening followed by full-text screening, will be independently conducted by two reviewers. A standardised data extraction template will be used, and qualitative data will be synthesised using thematic analysis. Quantitative data will be presented descriptively using graphs, diagrams, or tables.

**Ethics and dissemination:** As the study relied on publicly available research, institutional review board (IRB) approval is not required. Findings will be disseminated through peer-reviewed publications and conference presentations.

**Strengths and limitations of this study:**

- **Comprehensive Scope:** This review will encompass a broad spectrum of E-Health interventions, providing a holistic understanding of current practices within Indigenous communities globally.
- **Cultural Evaluation:** By examining cultural appropriateness alongside efficacy, this review will offer deeper insights into how interventions align with Indigenous values, traditions, and health practices.
- **Framework-Driven Rigour:** The methodology, guided by PRISMA-P and Arksey and O’Malley’s framework, ensures a systematic, transparent, and reproducible research process.
- **Identification of Barriers:** In addition to highlighting effective interventions, this review will uncover challenges and obstacles to adopting E-Health technologies, offering valuable information for policymakers, health professionals, and community stakeholders.
- **Language Restrictions and exclusion of Grey literature:** The findings will be limited to English publications, potentially restricting the inclusion of relevant studies published in other languages. Grey literature will be excluded from this scoping review, which may result in the omission of relevant unpublished or non-peer-reviewed research.

## INTRODUCTION

According to a report published by the International Labor Organization (ILO) in 2020, there are about 476.6 million Indigenous Peoples in the world, most of whom live in Asia and the Pacific (70.5%), Africa (16.3%), Latin America and the Caribbean (11.5%), Northern America (1.6%), and Europe and Central Asia (0.1%). (1, 2) Indigenous Peoples, although comprising only 6% of the world’s population, experience high rates (19%) of extreme poverty. (1, 3) These statistics reflect the ongoing impact of colonial policies, including forced cultural assimilation and economic marginalisation, welfare dependency, and dispossession from family, culture, and land. (4–10) Despite this, Indigenous Peoples continue to demonstrate resilience and resourcefulness in preserving their cultural heritage and improving their well-being. (11)

The United Nations Declaration on the Rights of Indigenous Peoples asserts the right of Indigenous communities to achieve the highest possible level of physical and mental health. Yet, healthcare systems worldwide frequently do not meet this objective.(12) Indigenous People frequently face significant challenges in accessing clinically appropriate and culturally safe healthcare. With a majority of Indigenous Peoples globally living in geographically remote areas, with limited public investment in essential services and digital infrastructure, they face physical, structural, and racial barriers to inclusion in the formal economy. (3) This is a prominent public policy issue that governments have attempted to address. (13–15) For example, over 66% of Indigenous Australians and 44% of Indigenous Canadians live in remote areas. (14, 16, 17) Indigenous health knowledge systems and voices are marginalised within mainstream healthcare systems, leading to a lack of inclusion of traditional cultural worldviews and spiritual healing methods. (18) This neglect deprives Indigenous Peoples of culturally safe and holistic healthcare and reflects a considerable deficiency in cultural sensitivity within numerous health educational environments. (4)

E-Health technologies present an opportunity to leverage the strengths and resilience of Indigenous populations in addressing health disparities. The World Health Organization (WHO) highlights that improving health service accessibility and quality through digital technologies is essential to achieving the Sustainable Development Goals. (19)

E-Health encompasses digital health interventions, web-based interventions, telehealth interventions, mobile Health (mHealth), electronic health records (EHR), telemedicine, and online interventions. These technologies have the potential to improve patient outcomes, streamline the effectiveness of the healthcare systems, and improve healthcare delivery, especially for historically underserved populations and those living remotely (20–22)

While E-Health offers considerable promise, many challenges remain. Geographic remoteness and socioeconomic differences contribute to gaps in Indigenous people’s digital access, creating a “digital divide” defined as community-level disparities in access and use of online technologies. This digital divide exacerbates already-existing inequalities experienced by Indigenous people in employment, healthcare, education, and community development, which underpin poor health outcomes for many

Indigenous Peoples. (21, 23, 24) Such structural conditions are illustrative of failures by government health systems to deliver culturally safe and responsive services, including health information. Combined with the broader impacts of colonialism, government system deficits contribute to increased susceptibility to diseases and lower life expectancy.(3, 25) By way of example, acute rheumatic fever (ARF) is typically prevalent in countries where there is high poverty, poor hygiene, and poor living standards. (26) Yet in Australia, a high-income country, 89% of new ARF and rheumatic heart disease (RHD) diagnoses between 2015 and 2017 affected Aboriginal and Torres Strait Islander people under 25 living in rural and remote communities. (27) In New Zealand, Māori and Pacific Islanders made up 93% of hospital admissions for ARF between 2002 and 2016. Localised studies from northwest Ontario in Canada, similarly, indicate comparably high and underreported rates of ARF within Indigenous communities. (27)

E-health interventions are a viable option for strengthening accessible and culturally tailored healthcare. They could improve accessibility, enhance cultural safety, and empower Indigenous People to manage their health. (28) Electronic modalities, such messaging and online chat rooms, offer anonymous social support and reduce stigma around diseases like HIV. (29) Evidence shows that E-Health provides Indigenous patients opportunity to access private, secure healthcare via phones, offering anonymous support and avoiding unwanted in-person interactions. (30–32) These technologies also offer speed, anonymity, accessibility, affordability, and convenience, reducing cultural and physical barriers to healthcare engagement. (31, 33, 34)

Indigenous People globally have varied and distinct cultural and geographic circumstances that require customised E-Health solutions. Limited internet access in remote locations is one of the many barriers to successfully implementing E-Health programs. (14, 35) Connectivity issues hinder the efficient delivery of healthcare through electronic medical records (eMR), electronic databases for pathology testing and referral forms, and health promotion and education. Integrating technological advancements with culturally sensitive health education and intervention methods is critical, but little guidance exists on how to design and implement such approaches. (36–39)

Numerous studies across diverse population groups have shown the growing popularity of E-Health interventions, particularly online, for addressing a range of psychosocial and physical health challenges. These include pain management, (40) substance abuse, (41) mental health, (42) sexual health, (43) heart health, (44) cancer, (45) diabetes, (46) and overall health management (47) (47) like medication compliance and encouraging physical activity. (40–42, 44, 45) Several online psychosocial programs have been tailored to support the needs of families or caregivers, and several systematic reviews have evaluated the efficacy of interventions across general populations, demonstrating equivalency between e-mental health interventions and in-person therapy for conditions like depression, anxiety, and trauma-related disorders. (48–51) However, not all populations are benefiting equally. The digital divide exacerbates already-existing health inequalities related to race, socioeconomic, educational, and age disparities. (20) Despite growing research on E-Health interventions for Indigenous communities, significant gaps remain. (52, 53) Previous studies have concentrated on particular illnesses, health challenges, and technological approaches without a comprehensive examination of E-Health’s specific applicability to Indigenous peoples, including its cultural appropriateness and role in therapeutic relationships. (54)

This scoping review aims to systematically explore the landscape of E-Health interventions for Indigenous populations, identifying effective strategies and gaps in current research and practice. By documenting evidence of E-Health use with and for Indigenous communities globally, this study contributes to integrating Indigenous health considerations into global health initiatives and national policies in alignment with the UN Declaration on the Rights of Indigenous Peoples. (21) This work seeks to contribute to broader efforts to address health disparities and advances toward achieving health equity for all.

## METHODS AND ANALYSIS

The method section provides an exhaustive description of the protocol for conducting a scoping review of E-Health interventions in Indigenous communities worldwide. Our scoping review protocol follows the Preferred Reporting Items for Systematic Reviews and Meta-Analysis Protocols (PRISMA-P) (55) Guidelines and the methodological approach first proposed by Arksey and O’Malley. (56) During the review process, the following stages will be completed:

1. Formulating the research question
2. Search strategy and identify relevant studies
3. Selection of eligible studies
4. Data Extraction
5. Collating, Summarising, and Reporting the Results

As this paper focuses on Indigenous populations, the lived experience of Australian Aboriginal woman Tyra Yarran will guide this research. Tyra, who is from Bardi Jawi Country in Australia, has over 20 years of experience working with Aboriginal and Torres Strait Islander Health systems and health challenges in a variety of health settings. She will be involved in all the next stages of the project.

### Stage 1: Formulating the research question

The review process commenced with an explicit delineation of the study’s question, objectives, and scope. We employed the epidemiological method of asking the questions of “Who? What? And When? to formulate a research question. Collaborative iterative discussions with subject matter experts led to the consensus that the review would answer the question: “In the last ten years, what research has been performed regarding E-Health Interventions in Indigenous Communities Across the World?” It was decided collectively to keep the research question broad in scope, avoiding specific diseases or geographic limitations. To obtain a complete grasp of the present research landscape, we did a thorough evaluation of scoping and systematic literature studies before finalising our research question. This comprehensive review aims to find literature gaps and prevent work repetition while developing our research topics. Notably, researchers like Intahchomphoo C. *et al*. 2018 (13) and Moecke *et al*. 2023 (54) were crucial in exposing reoccurring themes and particular difficulties experienced by these populations. Combining these evaluations’ findings, we created focused research questions that address understudied facets of digital inclusion and health outcomes in Indigenous contexts. This allowed us to ensure that our contributions are unique and effectively address the gaps that have been identified. This deliberate choice intended to collect comprehensive evidence about E-Health interventions in various Indigenous communities.

This research question guided the development of the specific study aims:

a. To identify and map the existing literature on E-Health interventions in Indigenous communities worldwide.
b. To assess the types and characteristics of E-Health interventions targeted at Indigenous communities.
c. To summarise the identified literature’s key themes, findings, and research gaps.
d. To inform future research priorities and policy decisions related to E-Health interventions for Indigenous communities.

This initial stage ensured the review’s focus was clear and laid the groundwork for subsequent phases.

### Stage 2: Search strategy and identifying relevant studies

The search strategy’s design started with identifying the relevant databases and the key terms within those databases. A comprehensive search will be conducted across multiple databases, including PubMed, Scopus, Web of Science, CINAHL, and Medline (Ovid). The search incorporated keywords, boolean phrases, Subject Headings, and Medical Subject Headings (MeSH) terms about E-Health and Indigenous populations.

Table 1 provides an overview of the selected terms and rationale for choosing databases. The final search strategy was drafted in consultation with the research librarian at James Cook University. The search scope was from January 2013 to the end of December 2024. The search will encompass published sources. To find additional relevant studies, the review will also use citation snowballing. Before uploading the finished dataset into Covidence for the ensuing review stages, all search results will be exported to Endnote and deduplicated by the primary reviewer.

**Table 1:** Example of Search Strategy.

| Serial Number | Medline Terms | Results |
| --- | --- | --- |
| **Indigenous** |  |  |
| 1 | exp Indigenous peoples/ or exp "american indian or alaska native"/ or exp "australian aboriginal and torres strait islander peoples"/ or exp maori people/ or exp "native hawaiian or other pacific islander"/ | 36,465 |
| 2 | exp Health Services, Indigenous/ | 4,155 |
| 3 | exp Indigenous Canadians/ or exp Inuit/ or exp Indians, North American/ | 18,346 |
| 4 | Indigenous health services.mp. | 137 |
| 5 | (indigen* or aborigin* or tribe* or "first nation*" or "native born*" or "native people*" or native* or "health Indigenous service*" or "Indigenous health service*" or "native hawaiian*" or "native hawaiian or other pacific islander*" or "pacific island american*" or Maori* or "australian aborigin*" or "australian race*" or "australoid race*" or "torres strait islander*" or "North American Indian*" or "Alaskan Native*" or Navajo* or Pima* or " South American Indian*" or "Central American Indian*" or "central american amerind*" or "central american indian*" or "north american amerind*" or "north american indian*" or "south american amerind*" or "south american indian*" or "metis canadian*" or "alaska Indigenous people*" or "alaska native*" or "alaska's Indigenous people*" or "alaskan native*" or "naabeeho*" or "navaho*" or "akimel o'odham*" or "akimel o'otham*" or inuit*).mp. [mp=title, book title, abstract, original title, name of substance word, subject heading word, floating sub-heading word, keyword heading word, organism supplementary concept word, protocol supplementary concept word, rare disease supplementary concept word, unique identifier, synonyms, population supplementary concept word, anatomy supplementary concept word] | 302,008 |
| 6 | 1 or 2 or 3 or 4 or 5 | 310,450 |
| **E-Health** |  |  |
| 7 | exp telemedicine/ or exp telepathology/ or exp teleradiology/ or exp telerehabilitation/ | 45,430 |
| 8 | exp Mobile Applications/ or exp Cell Phone/ | 30,467 |
| 9 | Gamification/ or Mobile Applications/ | 11,834 |
| 10 | (Telemedicine* or "mobile health*" or "mobileHealth*" or "tele-referral*" or telehealth* or "virtual medicine" or "virtual-medicine" or ehealth or mhealth or "eHealth*" or "m-health*" or "mobile app*" or "portable electronic app*" or "portable software app*" or "smartphone app*" or "software app*" or cellphone* or "cell phone*" or "mobile phone*" or "mobile-phone*" or "tele health*" or "electronic health*" or "electronic-health*").mp. [mp=title, book title, abstract, original title, name of substance word, subject heading word, floating sub-heading word, keyword heading word, organism supplementary concept word, protocol supplementary concept word, rare disease supplementary concept word, unique identifier, synonyms, population supplementary concept word, anatomy supplementary concept word] | 121,550 |
| 11 | (smartphone* or "smart-phone*").mp. [mp=title, book title, abstract, original title, name of substance word, subject heading word, floating sub-heading word, keyword heading word, organism supplementary concept word, protocol supplementary concept word, rare disease supplementary concept word, unique identifier, synonyms, population supplementary concept word, anatomy supplementary concept word] | 18,620 |
| 12 | 7 or 8 or 9 or 10 or 11 | 137,466 |
| 13 | 6 and 12 | 1,134 |
| 14 | limit 15 to yr="2013 - 2023" | 890 |
| 15 | **limit 15 to yr="2018 - 2023"** | **630** |

### Stage 3: Selection of eligible studies

After exporting the data to Covidence, the team will start the title and abstract screening process. Studies included in this scoping review will follow the eligibility criteria outlined in Table 2. These criteria will be circulated amongst all the reviewers. To ensure the alignment of the team on the scope of articles to be included, the primary reviewer will assign each reviewer 10 of the total articles for title and abstract screening. The team will meet to address the discrepancies within those 10, if any, and consolidate the inclusion and exclusion criteria. After the primary debrief, two primary reviewers will screen all the titles and abstracts independently. The second stage of the review process includes full-text screening. Studies that fit the potential inclusion criteria will be retrieved in full text during this screening phase. Full-text studies that do not meet the inclusion criteria will be excluded, and the final report will include the primary justification for this decision.

**Table 2:** Inclusion and Exclusion criteria.

| Component | Inclusion Criteria | Exclusion Criteria |
| --- | --- | --- |
| Study focus | Studies focusing on E-Health interventions in Indigenous communities across the world. | Studies not directly related to E-health interventions. Studies with insufficient information or methodology. |
| Population | Indigenous communities | Non-indigenous communities |
| Setting | Worldwide | --- |
| Language | English | Non-English |
| Type study | Primary research articles, systematic reviews, and technical reports. | Grey literature, editorial, opinion pieces, book chapters and protocols. |
| Year of publication | January 2013-December 2024 | Published before January 2013 |

A pair of independent reviewers will extract data, and their inter-rater reliability will be assessed and discussed about the theme content. The final report will contain comprehensive documentation of the search results and a PRISMA flow diagram to visualise them. As previously, discrepancies will be resolved by consensus or a third reviewer.

### Stage 4: Data Extraction

A standardised data extraction template (Table 3) will be used to collect information on the following:

**Table 3:**
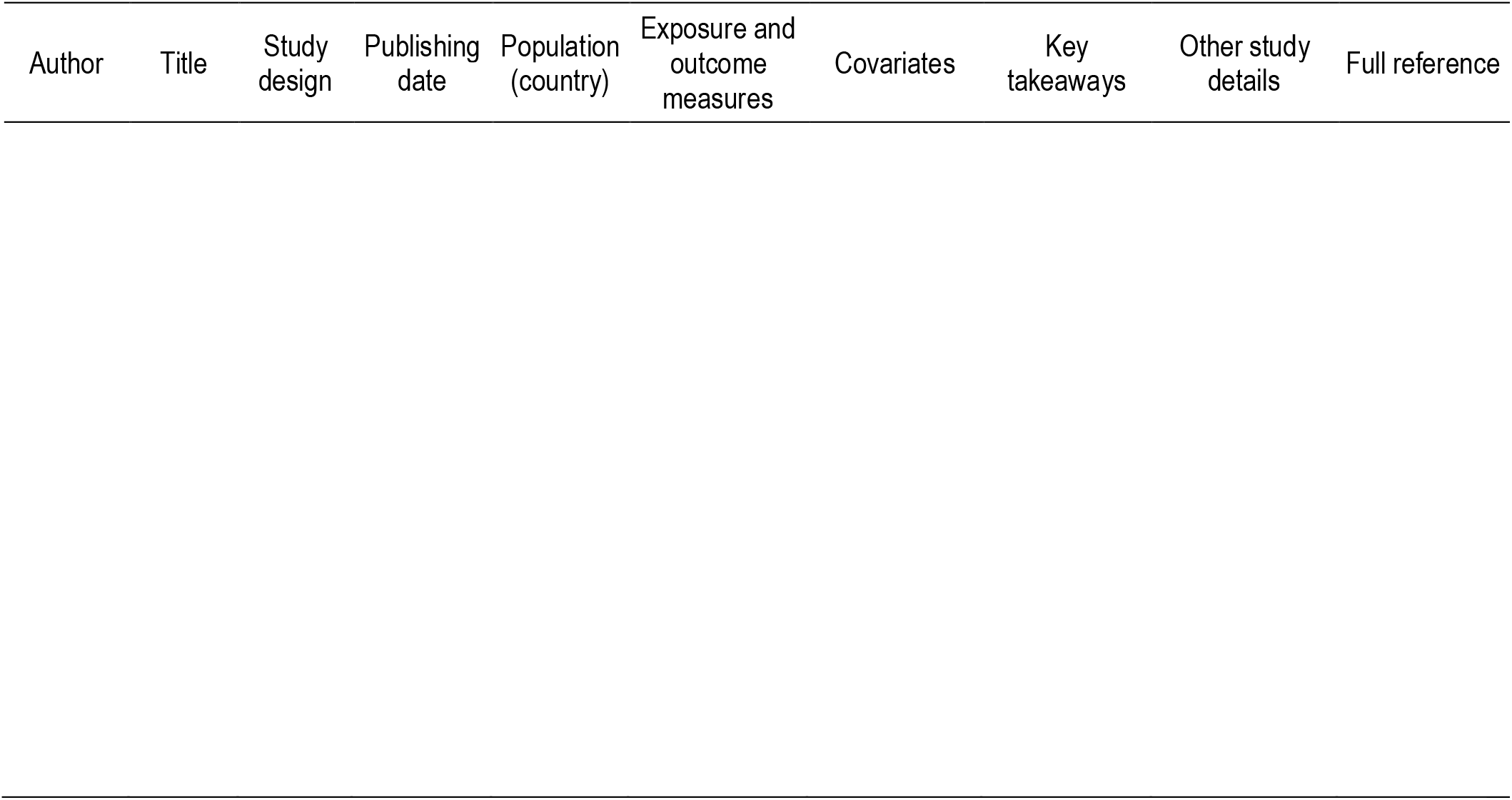
Data Extraction Instrument.

a. Study characteristics (e.g., authors, publication year)
b. Indigenous community demographics (e.g., location, size, culture, age mean/range, sex/female proportion)
c. E-Health interventions (e.g., type, purpose, technologies used, length)
d. Study methodology (e.g., study design, data collection methods)
e. Key findings and outcomes
f. Gaps or areas for future research

This template will later be expanded upon based on the characteristics of the findings. To ensure the process is validated, the data extraction template will be piloted in a random sample of three studies and then discussed within the team of authors. Edits will be made according to their suggestions. Two independent reviewers will then be assigned for data extraction. Discrepancies will be resolved by consensus or, if necessary, by a third reviewer. The full-text articles to be extracted will be stored in a shared library on EndNote and Google Drive. The Excel file for data extraction will be shared on Google Drive. A code book will be maintained to track the extractions done by the two reviewers.

We will use the Mixed Methods Appraisal Tool (MMAT) to do a risk of bias analysis and quality evaluation to maintain the integrity of our systematic review (57). Since the MMAT is intended to evaluate studies using mixed methods, qualitative, and quantitative approaches, it is perfect for reviews that cover a wide range of study designs. Our assessment process will include an initial screening to verify that studies fit the MMAT criteria. This will be followed by a thorough evaluation to appraise methodological quality along particular dimensions, including research design, data collecting, and data analysis. Every research will receive a score of ‘yes,’ ‘no,’ or ‘can’t tell.’ These results are combined to produce an overall quality rating for the methodology. (57)

### Stage 5: Collating, Summarising, and Reporting the Results

Data will be synthesised using thematic analysis. Quantitative data, if available, will be presented descriptively. The data will be presented graphically, diagrammatically, or tabularly.

Key themes and findings from the identified studies will be summarised and presented narratively. This narrative summary will describe how the findings relate to the study research question and objectives. The scoping review results will be reported following the Preferred Reporting Items for Systematic Reviews and Meta-Analyses Extension for Scoping Reviews (PRISMA-ScR) guidelines.

Descriptive statistics, if any, will be analysed using statistical software (SAS, R/R studio) or Excel. Data visualisation and graphs will be done using R/R Studio, Excel, or PowerPoint.

### Patient and public involvement

To ensure the cultural appropriateness and relevance of the findings, we will seek consultation and input from Indigenous communities, organisations, or experts in the field.

### Lived experience involvement

Tyra Yarran (nee Thomas), from Bardi Jawi Country is an Aboriginal Health Manager in West Australia (WA) Health with 20 years of experience in various health settings. Tyra has kindly guided the research team with her lived expertise, skills, and knowledge gained during her career in health. Tyra will be involved in all the stages of the project. In addition, Dr. Stephanie Topp, Dr. María Eugenia (Maru) Castellanos Reynosa, and Dr. Ginny Monterio have been instrumental in the development and success of this paper. Dr. Topp, a Professor of Global Health at James Cook University, specialises in health systems research and policy, with a focus on addressing healthcare inequities in low-resource settings. Dr. Castellanos Reynosa, a Senior Lecturer in Epidemiology and Communicable Disease Control, brings over 15 years of experience in infectious diseases research, particularly on tuberculosis, malaria, and HIV, with expertise in conducting epidemiological studies in Africa and Latin America. Dr. Ginny Monterio, with over 15 years of experience in change and transformation projects across Australia, the USA, India, and Colombia, focuses on helping organisations plan and implement sustainable change initiatives using evidence-based models and data analytics.

## ETHICS AND DISSEMINATION

As the study relied on publicly available research, IRB approval was not required. Findings will be disseminated through peer-reviewed publications and conference presentations.

## CONCLUSION

This scoping review protocol describes a comprehensive strategy to assess the breadth and depth of E-Health interventions in Indigenous communities worldwide. Recognising the unique health concerns of Indigenous populations, our research seeks to shed light on the landscape of E-Health solutions. We plan to investigate the efficacy, cultural appropriateness, and potential hurdles to adopting E-Health technology among Indigenous peoples using a careful research methodology guided by the PRISMA-P and inspired by Arksey and O’Malley’s methodological framework.

By emphasising the inclusion of E-Health interventions, this review aims not only to find and map current research but also to critically evaluate the qualities and effects of these studies. This review is expected to result in a greater knowledge of the types of E-Health interventions used, their effectiveness, and the extent to which they embrace cultural safety and foster therapeutic interactions within Indigenous communities. By examining these factors, we hope to gain useful insights into how E-Health might be used to reduce health disparities and advance health equity among Indigenous peoples.

By doing so, we aim to facilitate communication among researchers, healthcare practitioners, policymakers, and Indigenous communities themselves. Through this collaborative communication, we may aspire to design and deploy E-Health solutions that are not only technologically sophisticated but also culturally sensitive and inclusive, ensuring that the advantages of digital health breakthroughs are distributed equally.

Ultimately, this protocol sets the stage for a rigorous and meaningful exploration of E-Health interventions among Indigenous populations. It is a step towards acknowledging and addressing the digital divide, with the broader goal of ensuring that Indigenous communities worldwide can fully benefit from the advancements in healthcare technology, thereby moving closer to achieving global health equity.

## Data Availability

All data produced in the present study are available upon reasonable request to the authors

## Author’s contributions

Conception and design of the study: SS, MEC, GM, ST; Revision of the research question and protocol: RB, TY; Design of search strategy: SS, RB, MEC, TY, GM, ST; Data extraction: SS, RB, GM; Writing protocol: SS; Review and edit of the manuscript: RB, TY, MEC, GM, ST. All authors approved the final version of this protocol.

